# EEGBoostNet Ensemble for IoT-Based Brain–Computer Interface in Early Epileptic Seizure Detection

**DOI:** 10.1101/2025.10.03.25337233

**Authors:** Biplov Paneru

## Abstract

The goal of this study is seizure detection in four class datasets for different seizure stages in epileptic patients. An early notification system is created to simulate the behavior of the patient experiencing a seizure receiving emergency assistance from caregivers, and a dataset acquired from Mendeley is used to train different models. The proposed EEGNet-ET-XGB model’s (EEGBoostNet) effectiveness against hybrid deep learning models is demonstrated by the outcome of 95.88% and 94.41% mean accuracy on stratified cross validation. On the full dataset, Bi-GRU with attention, bidirectional LSTM-GRU models, and conventional ensemble techniques like XGBoost can all do remarkably well. Channel 9 data is the most important feature, according to the SHAP interpretability analysis, which is conducted on several models with the aid of SHAP plots. The IoT-BCI cloud modeling is adapted to make early notifications for emergency systems. This method is essential for categorizing different seizure types according to occurrences in order to provide early warning and for developing a home automation strategy that will help victims.

## 1. Introduction

As machine learning (ML) and artificial intelligence (AI) continue to advance, there is an increasing emphasis on using these technologies to improve clinical applications. Early disease diagnosis and prediction to facilitate early preventive intervention is a major objective in healthcare [1]. Researchers have focused a lot of emphasis on epileptic seizures, which are neurological disorders marked by particular patterns in electroencephalography (EEG) recordings. EEG signal feature extraction and pattern classification have been successfully accomplished using machine learning (ML) and deep learning (DL) approaches [2]. Recurrent seizures that result in unconsciousness or muscle spasms are a common neurological disorder called epilepsy, which has a major negative influence on a patient’s health, everyday activities, and general well-being [3].

Because EEG recordings offer vital information about the electrical activity of the brain, they are widely employed in epilepsy research [4]. EEG data analysis using ML and DL techniques has been the subject of numerous studies aimed at identifying and forecasting epileptic episodes [5,10,12]. More accurate seizure identification has been made possible by developments in EEG processing techniques, such as improved feature extraction methods, improved artifact removal, and improved time-frequency analysis [6]. About 50 million individuals worldwide suffer from epilepsy, which is one of the most common and dangerous neurological conditions, affecting 1% to 2% of the world’s population [7]. Given the severity of seizures and their risks, alongside the pressing demand for more accurate and advanced therapeutic approaches have been developed to identify seizure activity and deliver targeted electrical stimulation to help suppress seizures and reduce their frequency [9]. Brain-computer interface (BCI), sometimes known as brain-machine interface (BMI), is a buzzword in the field of neuroscience that involves converting human brain thoughts into a chip. These devices can be surgically implanted or placed externally. Such components enable the user to operate the actuators or detect input data via bilateral communication in order to complete the task [19].

This work solely focuses on development of hybrid models as well as ensemble, traditional ML models to find out the potentiality for deployment for aiding patient with epileptic seizure along with classifying it type and finally providing the aid as needed. The proposed research is divided into various section in which section I. is related to abstract, and general introduction along with related works. Section 2. Refers to methodology section in which various ML models are proposed and architecture proposed are shown. Section 3. Shows the results from the models as well as discussions from the results obtained finally. Comparing work with previous works and discussion on limitations of study. Final section deals with concluding the study describing its outcomes and potential application for real-world epilepsy related effects minimization with AI-powered technology. The work provides a framework for utilizing Medical Internet of Things (IoMT) for the purpose of early epilepsy detection and aiding patients.

### 1.1 Related works

In recent years, substantial progress has been made in applying machine learning (ML) techniques to predict and detect epileptic seizures using EEG signals. Rasheed et al. [1] presented a comprehensive review of state-of-the-art ML methods for early seizure prediction, highlighting existing research gaps, challenges, and potential future directions to enhance robustness and generalizability.

Several studies have utilized the UCI Epileptic Seizure Recognition dataset for classification tasks. For example, Kode et al. [2] evaluated multiple classifiers, including Extreme Gradient Boosting, TabNet, Random Forest, and a 1D Convolutional Neural Network (CNN), reporting accuracies of up to 99%, with their proposed 1D-CNN outperforming other models in accuracy, sensitivity, precision, and recall. Similarly, Kunekar et al. [3] compared conventional ML and deep learning (DL) approaches and found that Long Short-Term Memory (LSTM) networks delivered superior performance in seizure detection.

Vieira et al. [4] also proposed an LSTM-based method that achieved 97% validation accuracy while reducing features and channels, demonstrating that lightweight models leveraging Explainable AI (XAI) can achieve over 95% accuracy with only six features and five EEG channels, making them practical for mobile applications.

Moving beyond traditional architectures, Zhu et al. [5] introduced a multidimensional Transformer fused with recurrent networks (LSTM-GRU) to address the non-stationary and complex nature of EEG signals. Disli et al. [7] proposed a Continuous Wavelet Transform (CWT)-based depthwise CNN, which converted multi-channel EEG into image-like representations and achieved nearly 96% accuracy, outperforming earlier approaches without requiring additional feature extraction or classifiers. The potential of lightweight implementations for real-time use has also been explored; Tsanika et al. [8] developed a TinyML model using iEEG data, achieving up to 99% accuracy, highlighting the feasibility of deploying resource-efficient models in closed-loop neurostimulation devices.

Similarly, Abhisek et al. [9] explored fractal features and non-linear dynamics, achieving 100% accuracy in classifying focal versus non-focal EEG signals in the Bern-Barcelona dataset and outperforming state-of-the-art methods in interictal and preictal state detection. Brari et al. [10] introduced a correlation dimension-based feature extraction method, delivering near-perfect classification accuracy with computational efficiency. Additionally, Rab et al. [11] integrated deep learning with domain knowledge—such as frequency bands, electrode locations, and temporal patterns—while introducing XAI4EEG, a SHAP-based framework designed to improve interpretability and reduce validation time for clinicians.

Architectural innovations combining CNNs and recurrent models have also proven highly effective. Mallick et al. [12] developed a hybrid architecture using 1D convolutional layers, bidirectional LSTM, GRU, and pooling layers, achieving up to 100% accuracy for binary seizure detection on the Bonn dataset and strong results in multi-class classification. Esmaeilpour et al. [13] focused on detecting preictal states with CNN-based feature extraction followed by ensemble methods such as random forests and support vector machines, obtaining a sensitivity of 90.76% on the CHB-MIT dataset. Meanwhile, privacy-preserving approaches have gained traction;

Suryakala et al. [14] introduced a Federated Machine Learning (FML) method that trained decentralized EEG data models, achieving high sensitivity (98.24%) and specificity (99.23%) while ensuring patient data confidentiality. Collectively, these studies illustrate the breadth of techniques—from conventional ML and deep learning architectures to explainable and privacy-preserving methods—that are advancing the reliability and practicality of EEG-based seizure detection.

To address challenges with reliable keyboard-BCI, Paneru et al. (2025) [18] proposed an EEG-based BMI system focused on accurately identifying voluntary keystrokes through right- and left-hand movements. Their methodology involved preprocessing EEG signals with band-pass filtering, segmentation into 22-electrode arrays, and refinement into event-related potential (ERP) windows, resulting in a 19×200 feature array classified into three categories: resting state, ‘d’ key press, and ‘l’ key press. Recognizing the limitations of conventional models, the authors employed a hybrid deep learning architecture—BiGRU-Attention—to interpret the EEG signals effectively.

Su et al. (2025) [22] introduced the Epileptic EEG Detection and Recognition Model based on Multiple Attention Mechanisms and Spatiotemporal Feature Fusion (MASF), aiming to enhance the accuracy of epilepsy detection using EEG signals. Recognizing the limitations of CNNs in capturing global dependencies and LSTMs in handling long sequences, the authors integrated a hybrid attention mechanism, Transformer encoder, and dot-product attention to extract both spatial and temporal dependencies directly from raw EEG data—eliminating the need for complex preprocessing or handcrafted feature extraction. The proposed MASF model achieved impressive accuracies of 94.19% on the CHB-MIT dataset and 72.50% on the Bonn University dataset under ten-fold cross-validation, highlighting its robustness and generalization capability.

Despite these advances, further research is needed to address diverse seizure occurrences and strengthen early detection frameworks. The development of the Internet of Medical Things (IoMT) has introduced promising assistive strategies to improve patient care and quality of life. While earlier works often evaluated ML frameworks using only binary and four-class classifications [5], research has lagged in building models for three-class detection. With the dataset [15], we aim to develop ML models capable of both three-class and four-class classification of seizure events. Ultimately, the IoMT framework holds the potential to automate patient room and environmental monitoring while also enabling real-time emergency alerts to caregivers, thereby bridging clinical effectiveness with practical deployment.

## 2. Methods and Materials

### 2.1 Dataset description

A preprocessed dataset from mendely source is collected and no exclusive filtering or further preprocessing methods are applied as data set was ML trainable ready [15]

### 2.2 Preprocessing and features extractions

The preprocessing of EEG data here in the pipeline begins by loading raw .edf files for six patients (Patient 10 through 15), with each patient’s dataset consisting of multiple EEG recordings. The EEG files are read using the mne library with preload=True to allow in-memory processing. For consistent channel configuration, certain channels are either dropped or reordered depending on the patient ID to ensure uniformity across all recordings.

Once the data is loaded, seizure annotations are extracted using a custom Seizure_times module. Each seizure’s start time and duration are translated from real-time to sample indices using a sampling rate of 500 Hz. Based on these indices, seizure segments are sliced from the raw EEG matrix. These seizure segments are stored separately into three categories—CPS, electrographic, and nociceptive—depending on the patient ID and specific file conditions.

To balance the dataset, normal (non-seizure) EEG segments of equivalent length are also extracted from the parts of the signal that do not overlap with any seizure events. These normal and seizure segments are then broken into 1-second epochs (i.e., 500 samples per channel) and stored as 3D arrays with shape (epochs, channels, samples).

Each class of EEG data is normalized independently by dividing all values by the maximum absolute value within that class, ensuring that data scales are consistent across inputs. The labeled data are stacked together into a single feature matrix x, with labels assigned as follows: 0 for normal, 1 for CPS, 2 for electrographic, and 3 for nociceptive seizures. The final dataset is split into training and testing sets using an 90-10 ratio, and all arrays are saved in both .npy (NumPy) and .mat (MATLAB) formats for flexibility in further analysis or model training. All features were relevant so none exclusion was there. The training.npy available [15] on data source was used to train the models and test.npy sets for evaluating the model performance. So, no exclusive data splitting was required.

**Fig 1:**
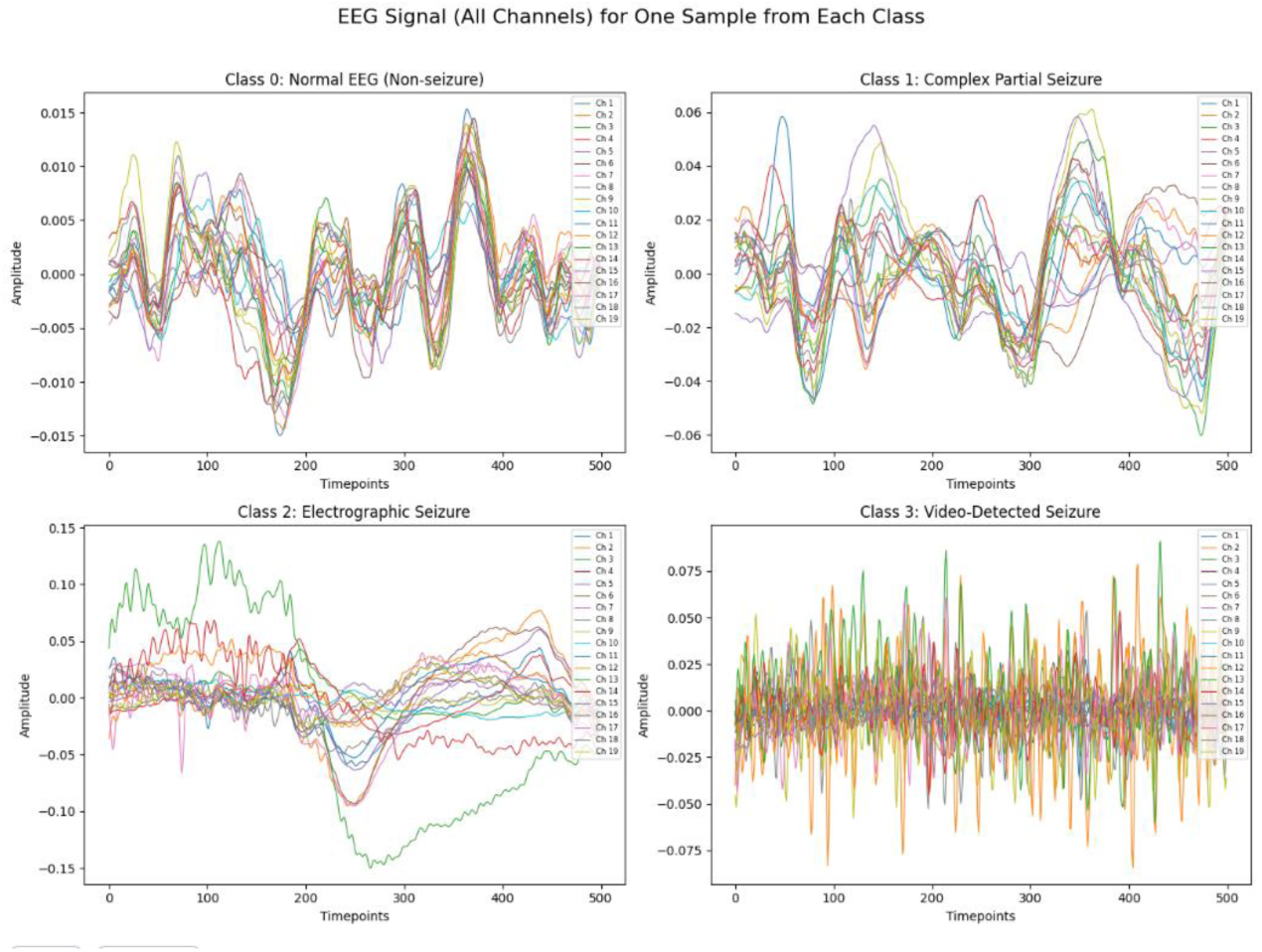
EEG signal variation plot for various events.

**Fig 2:**
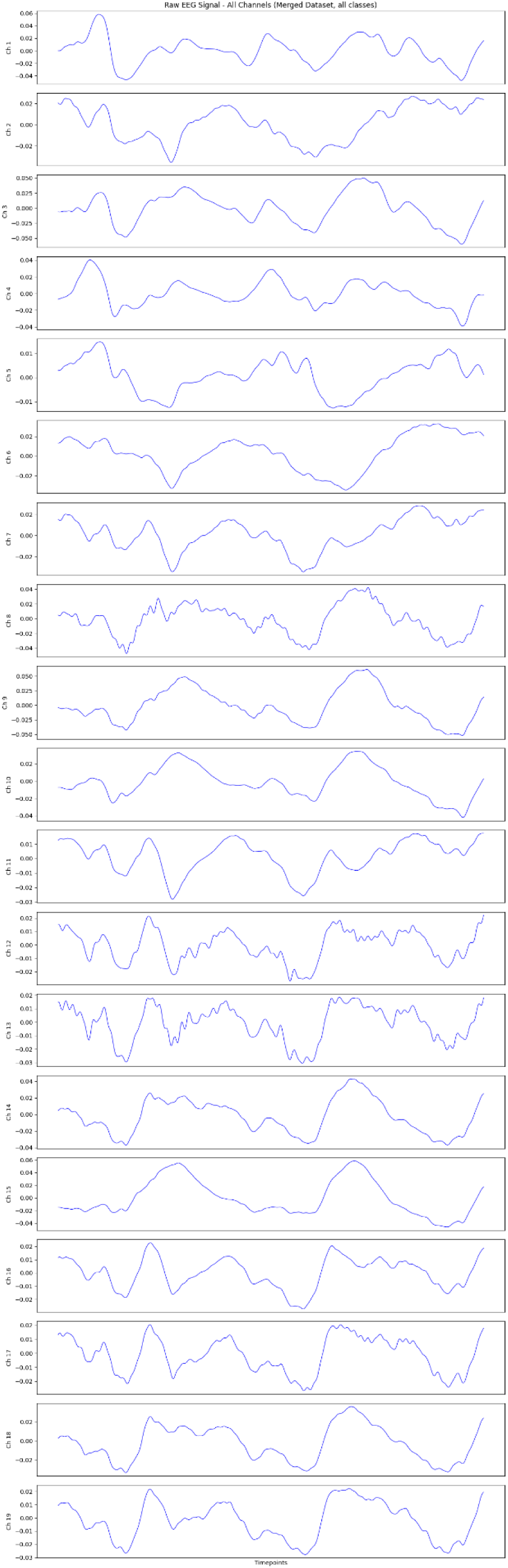
EEG data plot for all channels.

The EEG plots in the image illustrate representative signals across all 20 channels for one sample from each of the four classes. Class 0 (Normal EEG) shows low-amplitude, rhythmic waveforms without significant abnormalities, reflecting non-seizure brain activity. Class 1 (Complex Partial Seizure) exhibits moderate amplitude fluctuations and irregular patterns, indicating localized seizure onset. Class 2 (Electrographic Seizure) displays prominent amplitude shifts and gradual trends, characteristic of seizures detected solely through EEG. In contrast, Class 3 (Video-Detected Seizure) shows high-frequency noise-like patterns, possibly indicating seizure activity visible externally (e.g., motor movements) but less prominent in EEG amplitude.

#### Normalization

In this study, features for the stacked EEGNet-based model were extracted directly from the EEG signals through a deep convolutional neural network (EEGNet). The preprocessed EEG data, consisting of 19 channels and 500 time points per sample, were first standardized using z-score normalization, where the mean and standard deviation were computed across all training samples. Each input value was then transformed by subtracting the training mean and dividing by the training standard deviation, ensuring that the signals had zero mean and unit variance. This normalization not only stabilizes training for the deep network but also ensures that the test data are scaled consistently [21]. This is standard score (z-score) normalization which equations are given in equation 1, 2, 3 and equation 4.

#### Equations

For each element *x_i,j,k,l_* in the 4D tensor *x* ∈ ℝ^*N*×*C*×*T*×1^(samples × channels × timepoints × 1):

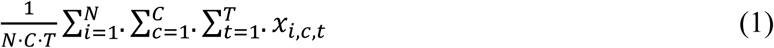

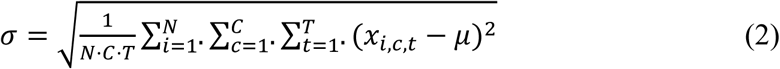

Then, normalized training data:

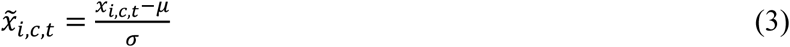

And test data is normalized using the training mean & std:

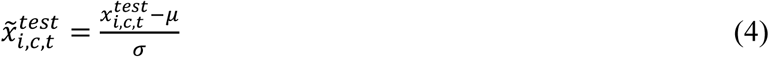

This ensures the test data is scaled consistently with the training data, which is standard practice in ML and deep learning.

### 2.3 EEG 10-20 System

The standard 10–20 electrode system is a globally recognized technique for applying electrodes to the scalp in order to capture electroencephalogram (EEG) information. The term “10-20” describes the electrode placement, which occurs at intervals of 10% or 20% of the total distance between particular anatomical landmarks on the skull, including the preauricular points (sides), inion (back), and nasion (front).

**Fig 3:**
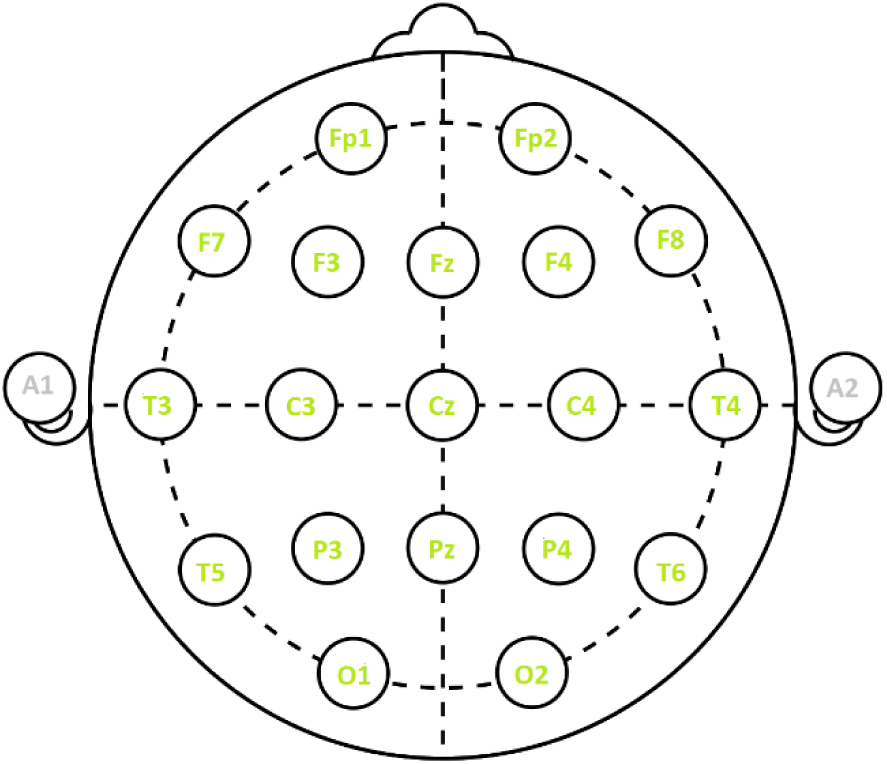
10-20 Electrode system.

Electrodes are labeled with numbers that indicate the placements of the left (odd numbers) or right (even numbers) hemispheres, and letters that indicate the region of the brain (F for frontal, T for temporal, P for parietal, O for occipital, and C for central). The 10-20 system is frequently used in neurophysiology, clinical diagnosis, and research to record brain rhythms, identify anomalies like epilepsy, and investigate cognitive processes.

### 2.4 Seizure directory

The seizure dictionaries (seizures_10 to seizures_15) served as crucial metadata for the EEG data preprocessing pipeline. Each dictionary corresponded to a specific patient and contains information about the onset time and duration of seizure events recorded in different EEG sessions. The structure of the data—formatted as {record_number: [(hour, minute, second, duration), …]}— allows precise temporal localization of seizure episodes within each recording file. During preprocessing, these timestamps are converted into sample indices using the known sampling rate (500 Hz), enabling the exact extraction of seizure segments from the raw EEG signals. This time-to-index conversion is essential for segmenting the data into meaningful 1-second windows for model training. These seizure labels also ensure that the data is categorized correctly into seizure and non-seizure classes (e.g., CPS, electrographic, nociceptive), which is critical for training accurate and clinically useful machine learning models. Without these annotated seizure dictionaries, it would not be possible to align the EEG data with the actual clinical events, thus significantly compromising the validity and effectiveness of the downstream classification tasks.

#### i. Bi-Directional LSTM GRU

This model combines Bi-Directional Long Short-Term Memory (LSTM) and Gated Recurrent Unit (GRU) layers to capture temporal dependencies in sequence data from both past and future contexts. The bi-directional setup allows the network to understand patterns from forward and backward sequences, improving performance on time-series tasks like EEG signal classification.

#### ii. Extra Trees

Extra Trees (Extremely Randomized Trees) is an ensemble learning method that builds multiple decision trees using random splits of features and thresholds. Unlike Random Forests, Extra Trees inject more randomness, often leading to faster training and sometimes better generalization. It is effective for tabular data classification and regression tasks.

#### iii. Stacked Ensembled model with EEGNet

EEGNet is a compact convolutional neural network architecture specifically designed for EEG signal processing. It efficiently extracts spatial and temporal features from EEG data using depthwise and separable convolutions, making it well-suited for brain-computer interface (BCI) applications with limited training data and low computational cost [16].

After training, the softmax output probabilities from the final dense layer of EEGNet were used as high-level features representing the discriminative patterns learned from the EEG signals. These probability features were subsequently standardized again using column-wise z-score normalization before being passed to the tree-based base learner (ExtraTreesClassifier) and meta-learner (XGBClassifier) in the stacking framework. This two-stage feature extraction and normalization approach allows the model to leverage both deep representations of EEG patterns and robust ensemble learning for improved classification performance.

The proposed system employs a stacked ensemble model combining EEGNet, ExtraTrees, and XGBoost to detect epileptic events from EEG signals. EEGNet, a compact convolutional neural network, extracts deep spatiotemporal features from raw EEG time-series, capturing relevant neural patterns. These learned features are then standardized and passed to ExtraTrees, a tree-based base learner that models complex relationships and reduces variance. Finally, XGBoost acts as a meta-learner, integrating the predictions from ExtraTrees to produce robust final classifications. This hybrid architecture leverages both deep learning and classical machine learning strengths, improving detection accuracy and reliability for real-time BCI applications in an IoT-enabled healthcare environment.

**Fig 4:**
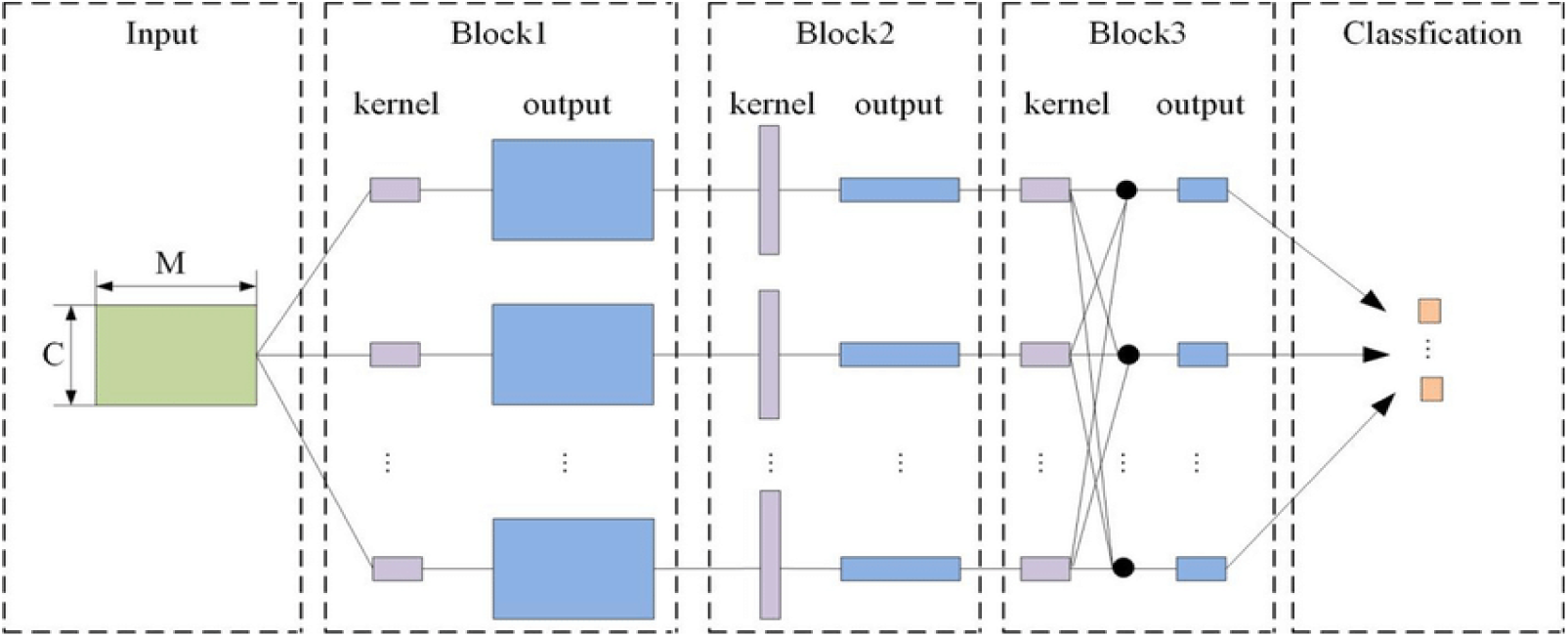
EEGNet architecture [16].

**Fig:**
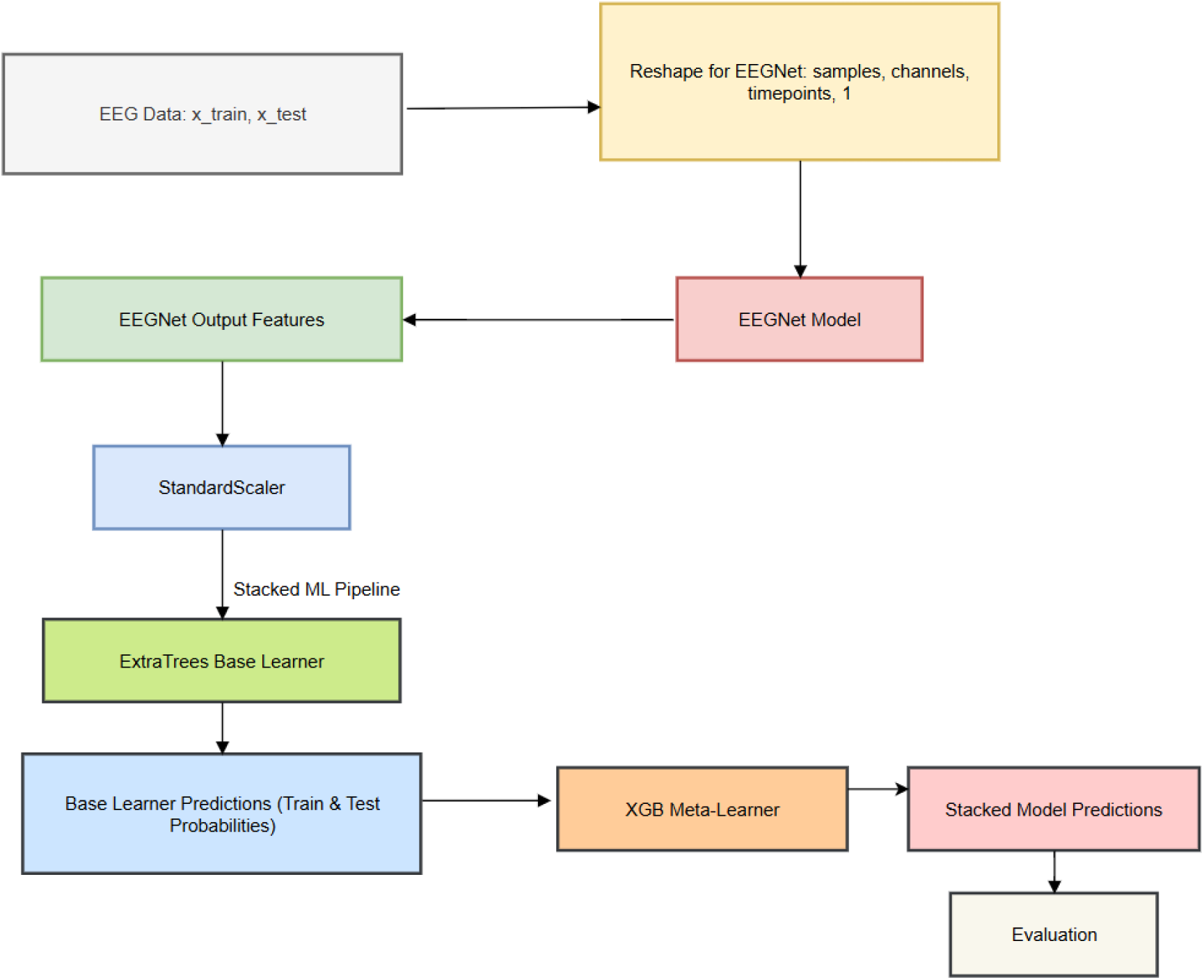
Stacked model pipeline architecture diagram.

#### iv. Bi-Directional GRU with Attention

This model enhances a bi-directional Gated Recurrent Unit (GRU) network by incorporating an attention mechanism that allows the model to focus on important time steps in the sequence. The attention layer improves interpretability and performance by weighting relevant parts of the input, which is particularly useful for complex sequential data such as EEG signals [18].

### 2.5 Proposed Workflow

First, models are feature extracted as preprocessing was already done on dataset they are trained and evaluated with various evaluation metrics. In order to find out the best performance, the nested cross-validation technique is applied. This enhanced the model development as well as ensured the model’s performance isn’t compromised on different folds of the dataset. With this cross-validation technique, model performance on the entire dataset can be rigorously tested. Then, the trained model is proposed to be deployed on the proposed IoMT system-based concept to aid patients.

**Fig 5:**
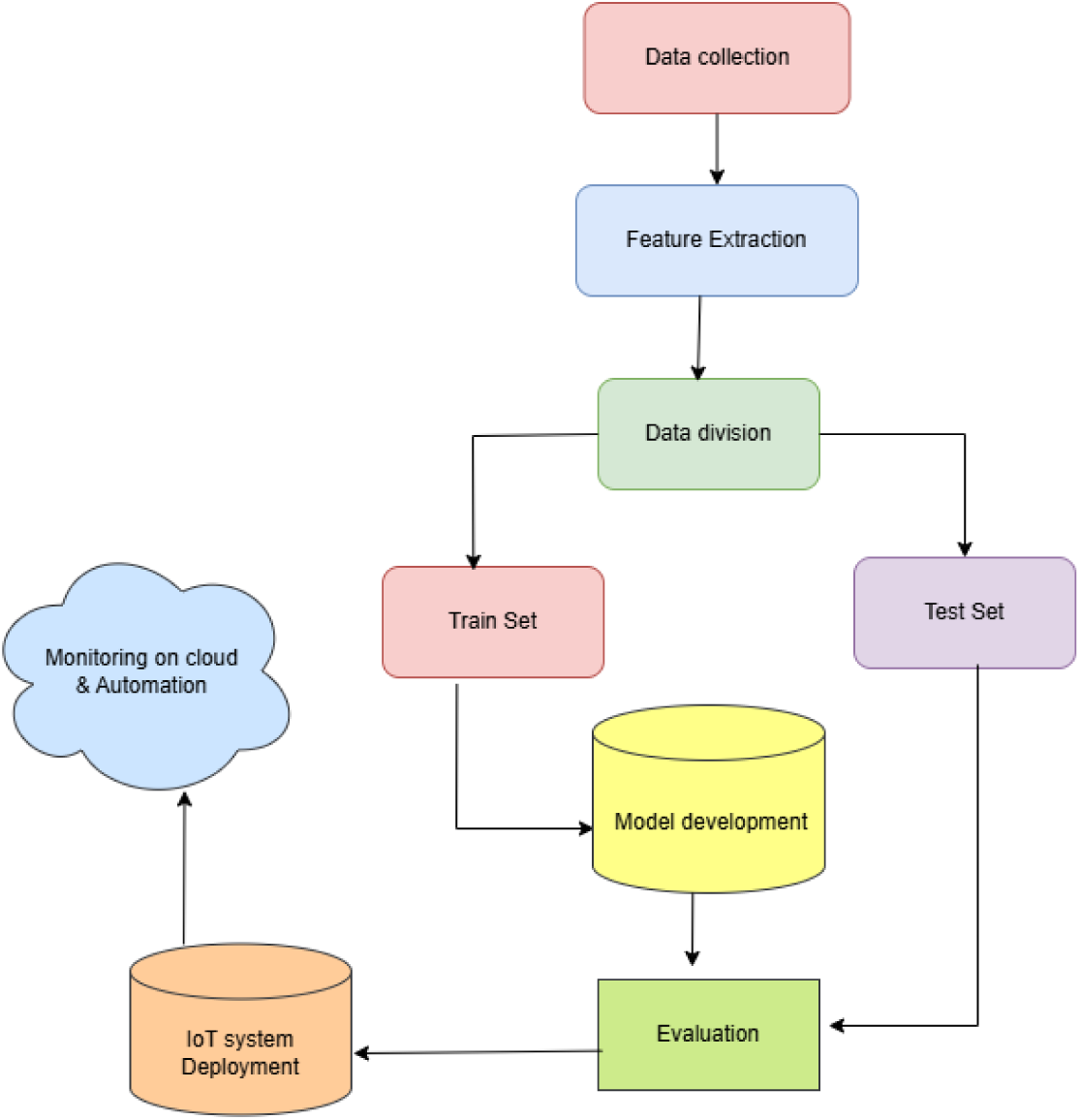
Proposed modeling concept.

The various ML models are trained to make seizure detection models. Two hybrid models are conceptualized that performed well beyond EEGNet a common model in ML field. The dataset includes EEG segments (each of size 19 channels × 500 time points, i.e., 1 second at 500 Hz), and each segment is labeled with one of the following: The dataset labels are tabulated in table 1.

**Table 1:**
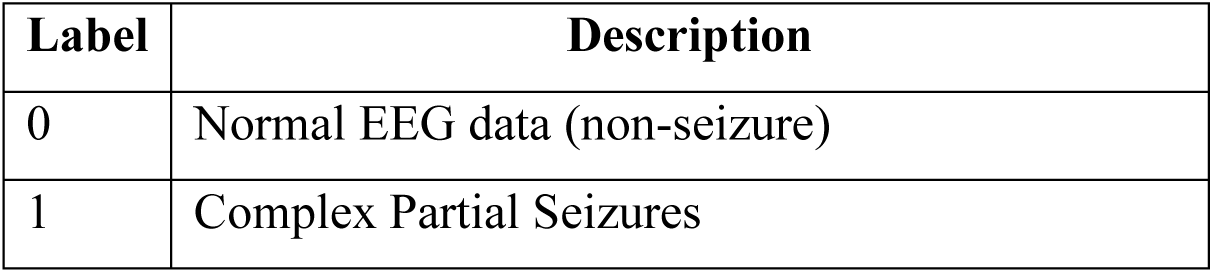
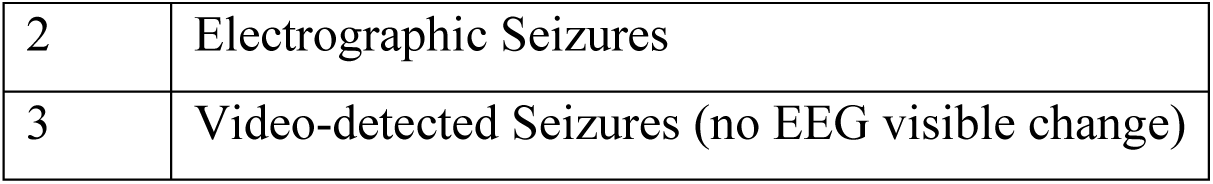
Labels on dataset.

| Label | Description |
| --- | --- |
| 0 | Normal EEG data (non-seizure) |
| 1 | Complex Partial Seizures |
| 2 | Electrographic Seizures |
| 3 | Video-detected Seizures (no EEG visible change) |

### 2.6 IoMT-based approach (IoT and EEG interaction for aid)

The Internet of Things with cloud databases can be used to record the data and provide feedback as a brain machine interface or early aiding for patients. BCIs allow direct communication between the brain and external devices, providing a lifeline for people with significant physical limitations. Incorporating IoT concepts may improve BCI efficacy. The integration of BCI with IoT technology offers distinct advantages. BCIs can be linked to the IoT framework to provide a more adaptable and comprehensive communication and control system [20].

In the proposed system (Figure 6), real-time EEG signals are acquired from the patient via a headset connected to a microcontroller, which continuously streams data to an IoT platform. The data is processed by the pre-trained seizure detection model, enabling early identification of potential epileptic events. Upon detection, alerts such as emails, SMS, or other notifications can be automatically sent to caregivers or medical personnel to provide timely assistance.

**Fig 6.**
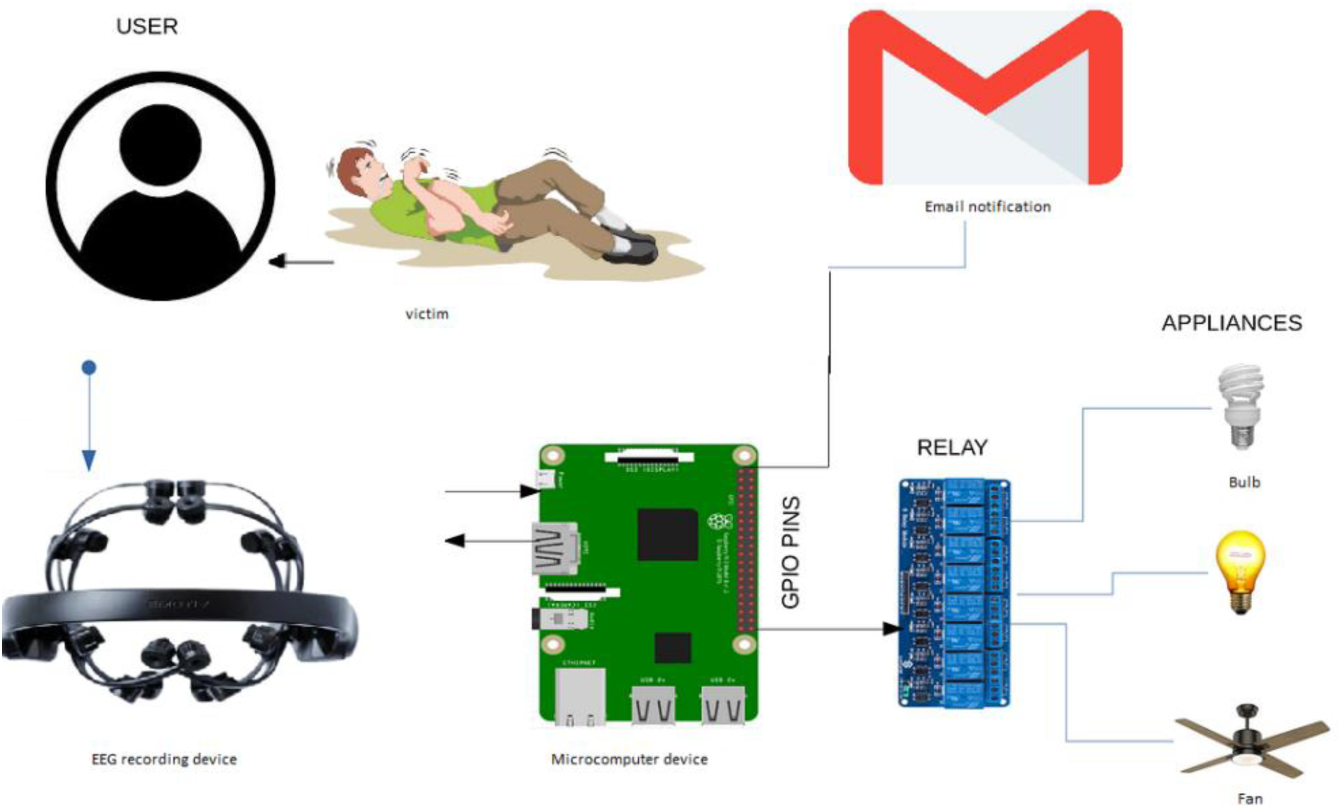
(a): Proposed system concept.

Additionally, the system allows automation of the patient’s environment: connected appliances such as fans, lights, or other devices can be triggered via relay circuits to adjust room conditions safely during a seizure. The model deployed on the IoT platform supports both real-time inference and incremental learning, allowing newly acquired EEG data to be used for retraining, thereby improving predictive performance over time.

**Fig 6.**
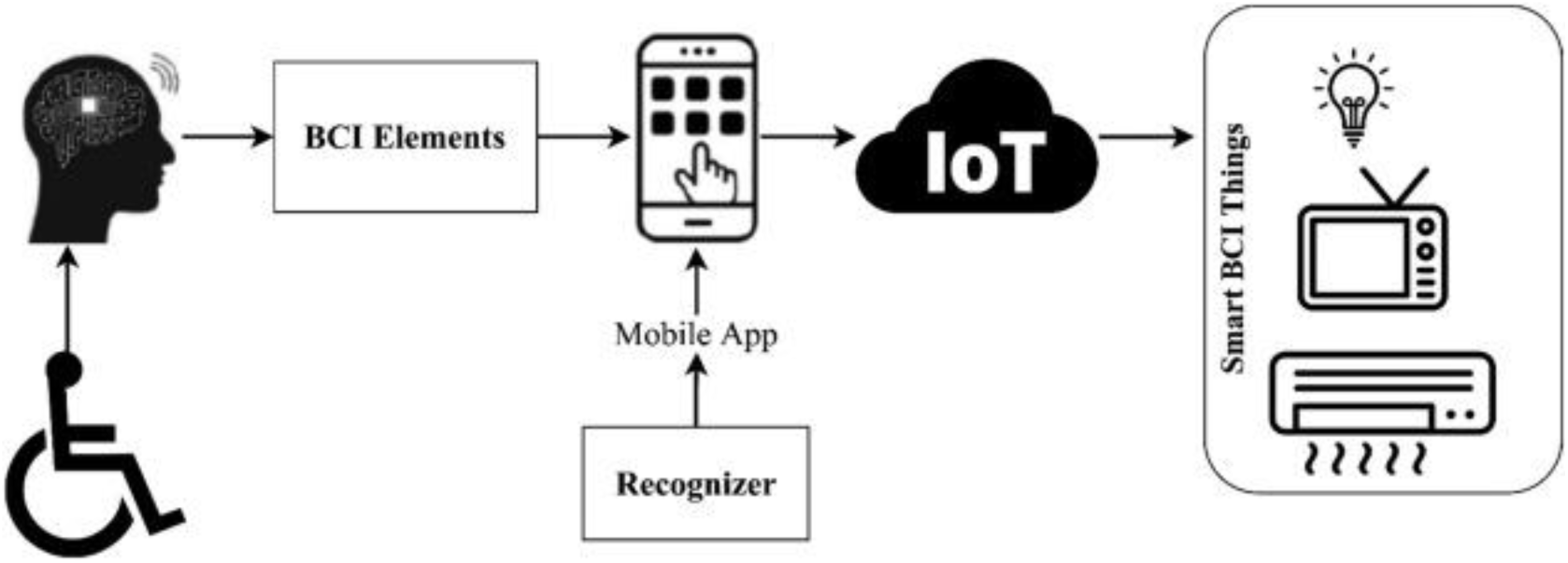
(b): IoT enabled BCI [19].

This integration of EEG headset, microcontroller, IoT infrastructure, and predictive modeling ensures continuous monitoring, timely alerts, and automated support for epileptic patients, enhancing safety and response efficiency.

With this concept, early aid can be made available to facilitate the victim and prevent the deterioration of his/her health further. With early notification, the caretaker can be called early to aid the patient.

### 2.7 Evaluation metrics

Accuracy, precision, recall and F1-score: Accuracy measures the overall correctness of a classification model by calculating the proportion of total predictions that are correct. Precision evaluates how many of the positive predictions made by the model are actually correct, reflecting the model’s ability to avoid false positives. Recall (also known as sensitivity) measures the model’s ability to identify all actual positive cases by finding the proportion of true positives detected out of all real positives, thus indicating how well the model prevents false negatives. The F1-score is the harmonic mean of precision and recall, providing a balanced metric that combines both aspects to obtain one performance measure, especially useful when the class distribution is imbalanced [17].

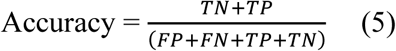

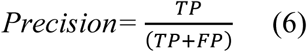

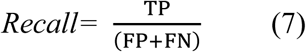

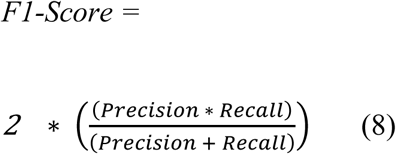

### 2.8 SHAP analysis

First, after training the Random Forest classifier on the scaled and flattened EEG data, we have used the SHAP (SHapley Additive exPlanations) library to interpret the model’s predictions on the testing dataset. We start by creating a TreeExplainer object specific to the trained Random Forest model, which efficiently calculates SHAP values. These SHAP values quantify the contribution of each feature to the prediction for each test sample. Since the problem is multiclass classification, the SHAP values are computed separately for each class, resulting in a list of arrays—one per class.

To visualize the impact of features for a chosen class, we have generated a SHAP summary plot, which displays the distribution and magnitude of SHAP values across all features and samples, highlighting how features influence model output. Additionally, a SHAP bar plot is created, showing the mean absolute SHAP values per feature as a measure of overall feature importance. Both plots provide intuitive insights into which features drive the model’s decisions. Because the input features correspond to flattened time-series data across multiple EEG channels and timepoints, we also reshape the model’s built-in feature importances back into the original two-dimensional shape (channels × timepoints).

Finally, by averaging feature importance values over timepoints, we produce a bar plot summarizing the relative importance of each EEG channel. This combined visualization approach helps understand both granular and aggregated contributions of the input data to the classification results.

## 3. Results and Discussions

In this section, the performance of several models is compared and reviewed, along with SHAP explanations for identifying relevant features in prediction. The possibility for incorporating a model for early aid for victims’ conceptual framework is also reviewed here, as are the study’s limitations and future directions.

### 3.1 Models performance evaluations

#### a. Bi-Directional GRU with attention method

The confusion matrix for Bi-Directional GRU with attention approach model obtained a training Accuracy of 89% and testing accuracy of 84%.

The confusion matrix plot in figure 7 is displayed as seen in figure in which for class ‘0’ 378 instances or testing feature set were correctly classified for class ‘0’ similarly, for class ‘1’ 230 instances were correctly classified as class ‘1’ and ‘43’ instances for class ‘2’.

**Fig 7.**
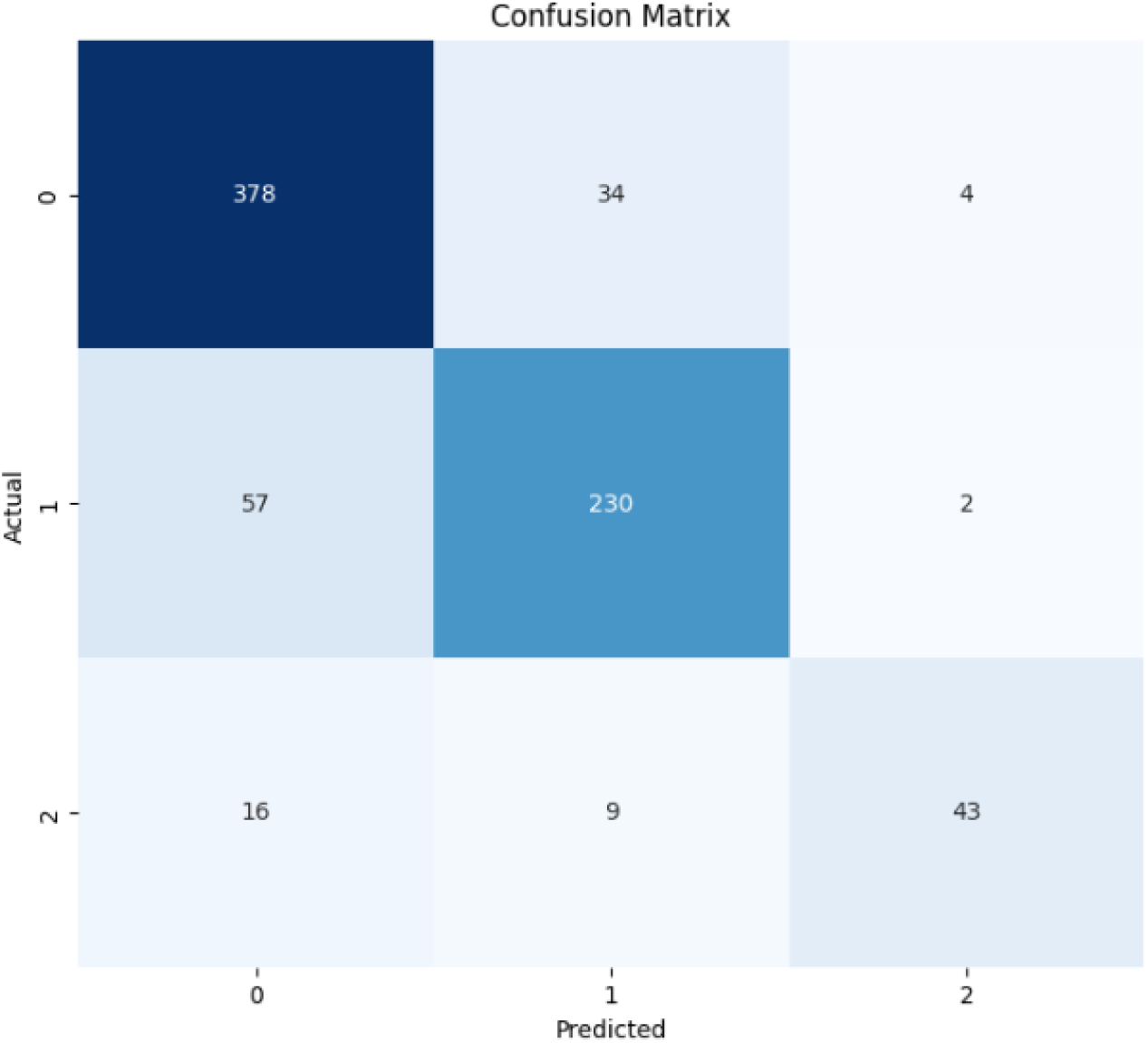
Confusion matrix for Bi-directional GRU with attention.

The model shown good development over epochs with improvements. The training history plots illustrate how the model learns over approximately 45 epochs. In the left plot showing accuracy, both training and validation accuracy rise sharply in the initial epochs, indicating that the model quickly captures key patterns in the data. The training accuracy stabilizes around 0.88–0.90, while the validation accuracy plateaus slightly lower, around 0.83–0.85, suggesting that the model is fitting well but retains a modest gap between training and validation performance. In the right plot, depicting loss curves, both training and validation loss decrease steeply at first and then flatten out, maintaining similar levels from about epoch 10 onward as shown in figure 8. The close alignment between training and validation loss throughout training suggests that the model generalizes reasonably well and does not suffer from significant overfitting or underfitting. Overall, these plots indicate a stable and effective training process, achieving good performance while maintaining consistency between training and validation results.

**Fig 8.**
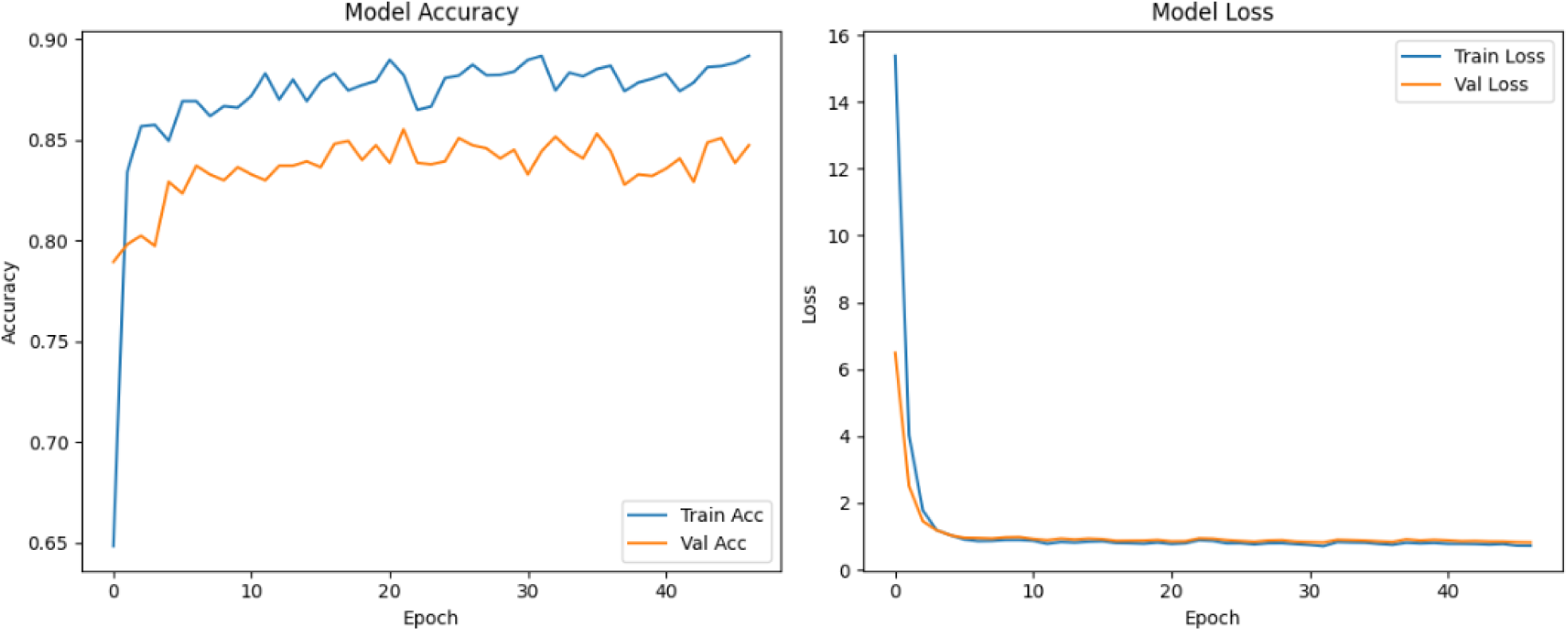
Training and loss history plot.

### b. EEGNet stacked with XGB + ET + RF

.The proposed system employs a stacked ensemble model combining EEGNet, ExtraTrees, and XGBoost to detect epileptic events from EEG signals that performed exceptionally well on dataset with stacked version.

The model EEGNet obtained a test accuracy of 95.98% including train accuracy of 99.49%. The confusion matrix plot is displayed as seen in figure 10 in which for class ‘0’ 400 instances or testing feature set were correctly classified for class ‘0’ similarly, for class ‘1’ 274 instances were correctly classified as class ‘1’ and ‘68’ instances for class ‘2’.

**Fig 10:**
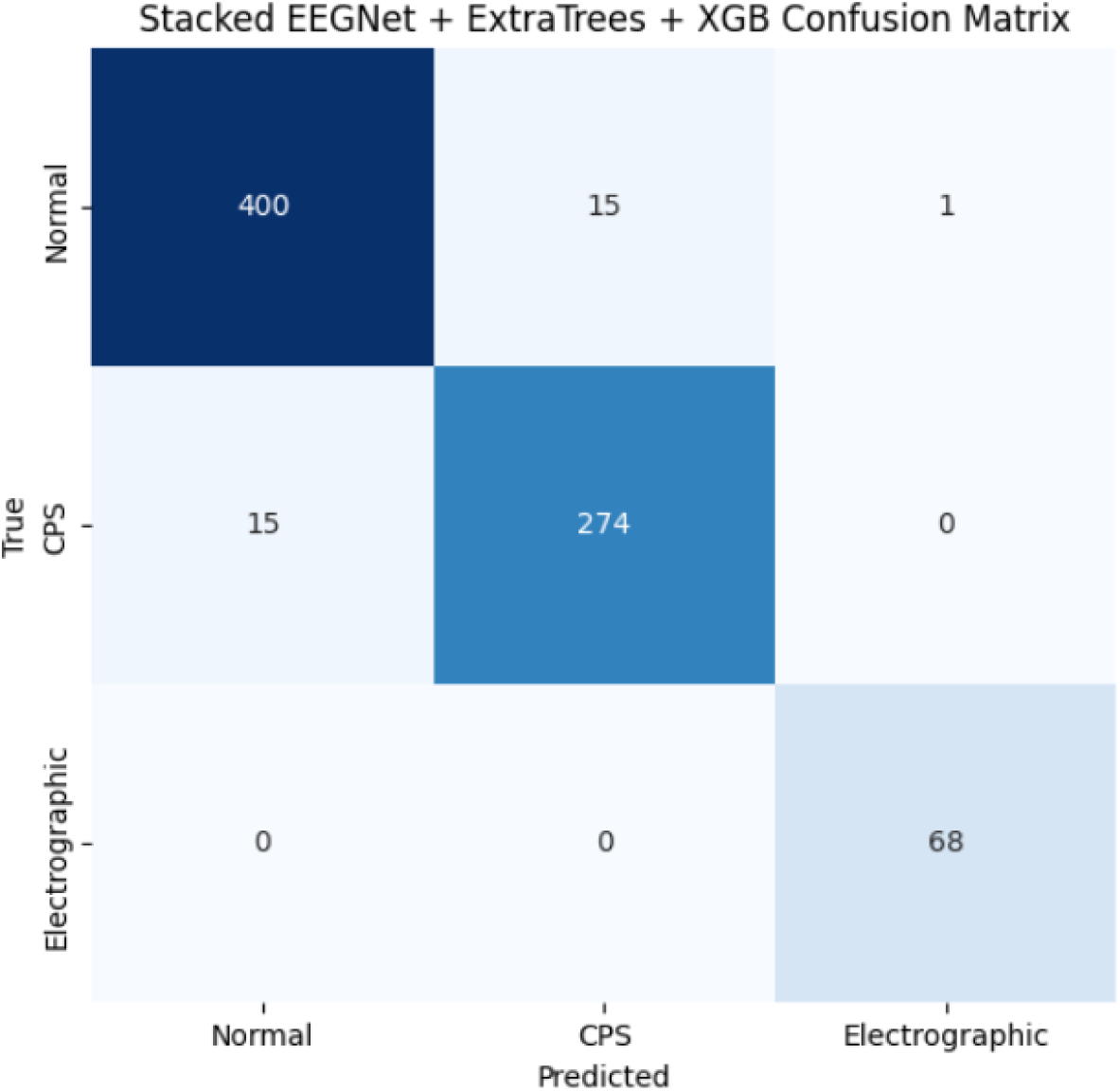
Confusion matrix plot for model.

#### 5-Fold stratified cross validation result

The hybrid EEGNet + ExtraTrees + XGB stacked model demonstrates strong performance in EEG classification, as shown by the 5-fold stratified cross-validation. In this setup, EEGNet first extracts temporal and spatial features from the EEG signals, normalizing the data to ensure consistent scale across channels and time points. These extracted features, in the form of softmax probabilities, are then fed into tree-based ensemble models—ExtraTrees as a base learner and XGBoost as a meta-learner—which further capture nonlinear patterns and complex interactions among the features. Across the five folds, the model achieved test accuracies of approximately 94– 95%, with a mean accuracy of 94.41% and a standard deviation of 0.88%, indicating both high performance and stability. This stratified cross-validation approach as shown in table 13 ensures that each fold maintains the same class distribution, which is particularly important for EEG datasets that often have imbalanced classes. The combination of deep feature extraction by EEGNet and robust classification by ensemble methods provides a significant advantage, as it leverages both learned representations from raw EEG data and the ensemble’s ability to reduce variance and improve generalization. This demonstrates that the hybrid model can reliably classify EEG patterns, which is crucial for applications such as seizure detection or cognitive state monitoring.

##### c. Extra Trees classifier results

The model Extra Trees classifier obtained a training Accuracy: 96.7% and testing Accuracy: 90% and confusion matrix is depicted in figure 11.

**Fig 11:**
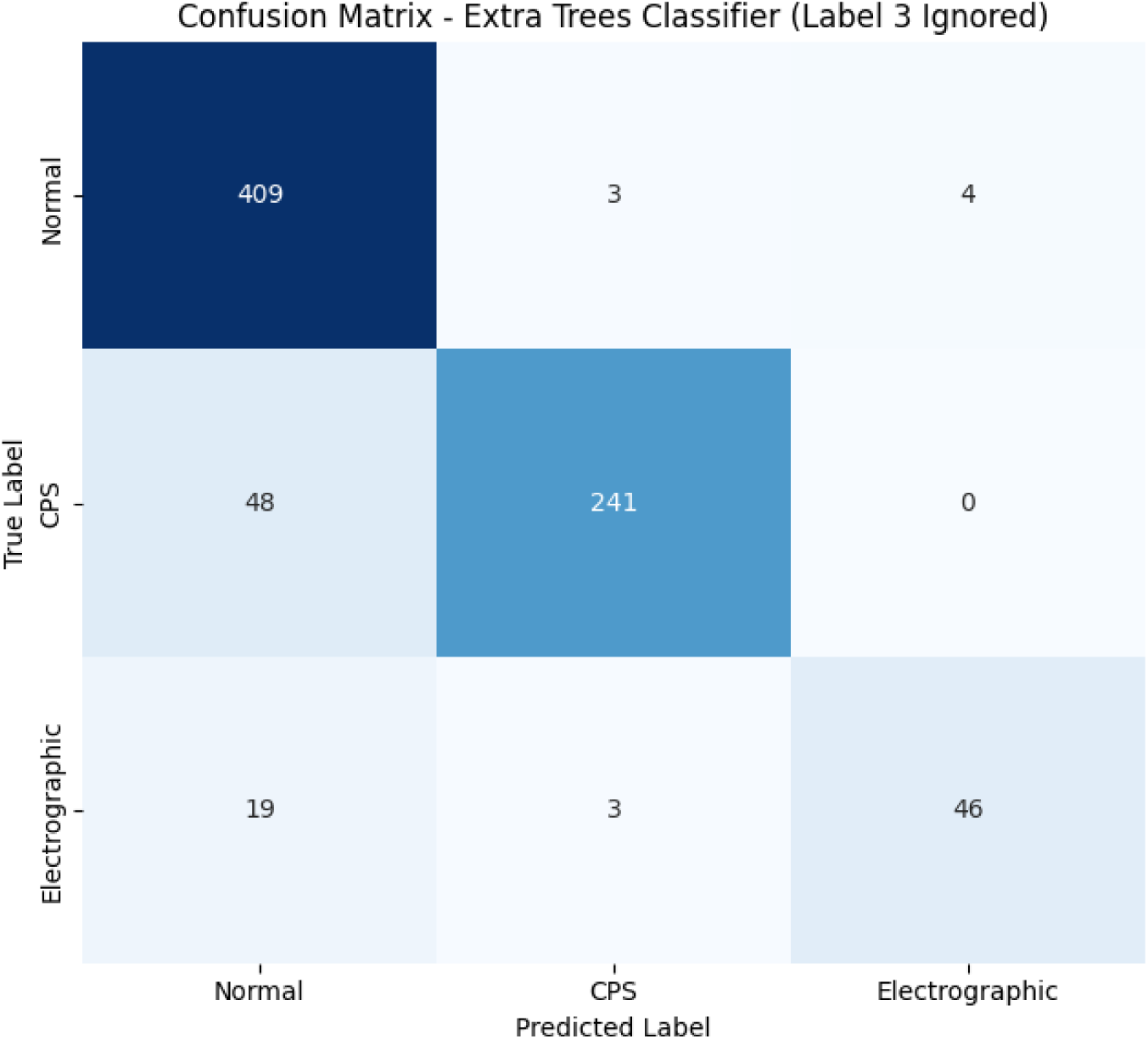
Confusion matrix plot for Extra Trees Classifier model.

##### d. Random Forest model

The model obtained a training accuracy of 100% as well as testing accuracy of: 0.88. The confusion matrix plot is displayed as seen in figure in which for class ‘0’ 404 instances or testing feature set were correctly classified for class ‘0’ similarly, for class ‘1’ 247 instances were correctly classified as class ‘1’ and ‘34’ instances for class ‘2’ as shown in figure 12.

**Fig 12:**
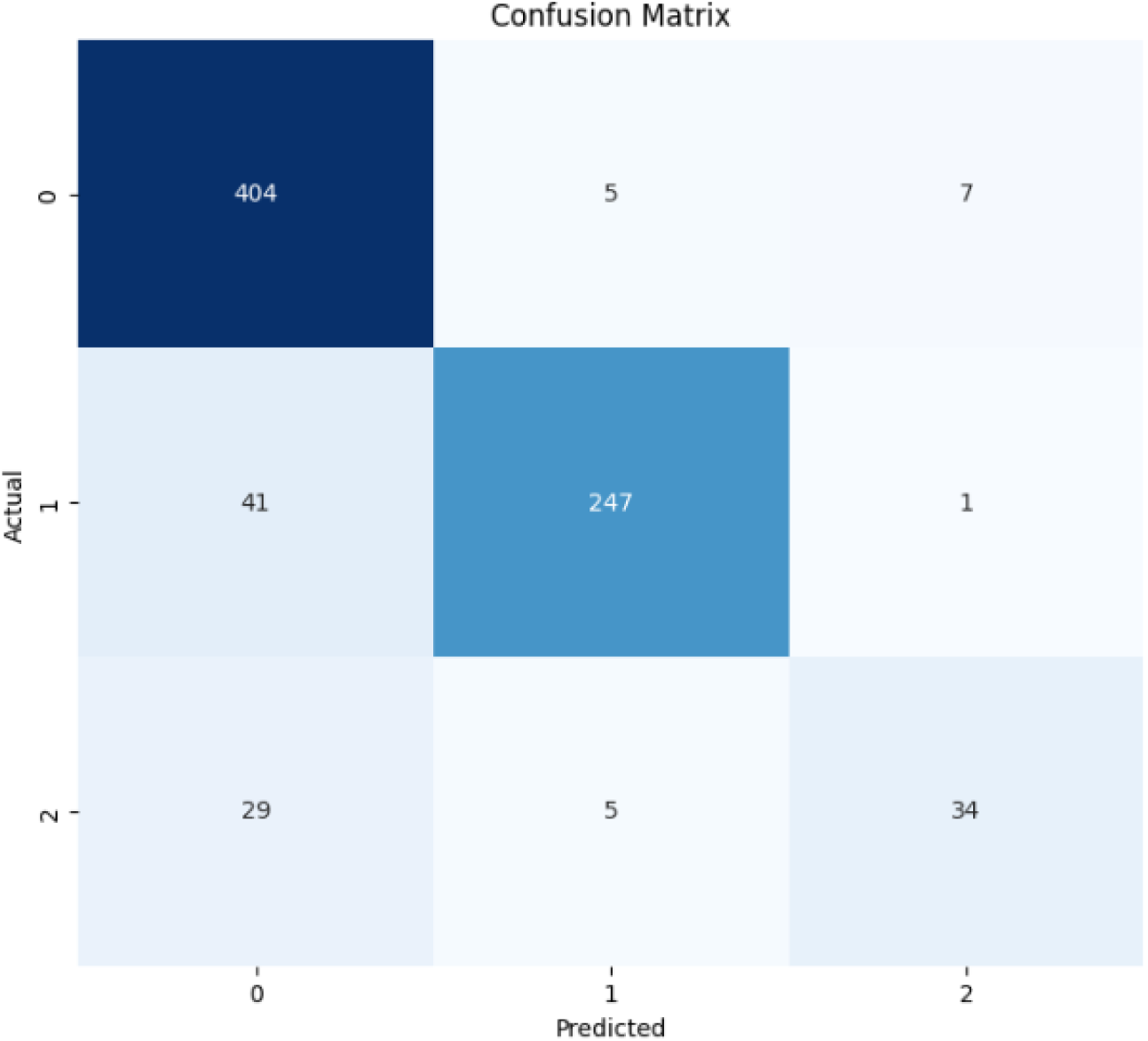
Random forest model confusion matrix plot.

##### e. Bi-Directional LSTM GRU

The model obtained a training Accuracy: 0.89 and testing accuracy of 0.84. The confusion matrix plot is displayed as seen in figure in which for class ‘0’ 371 instances or testing feature set were correctly classified for class ‘0’ similarly, for class ‘1’ 236 instances were correctly classified as class ‘1’ and ‘41’ instances for class ‘2’ as shown in figure 13.

**Fig 13:**
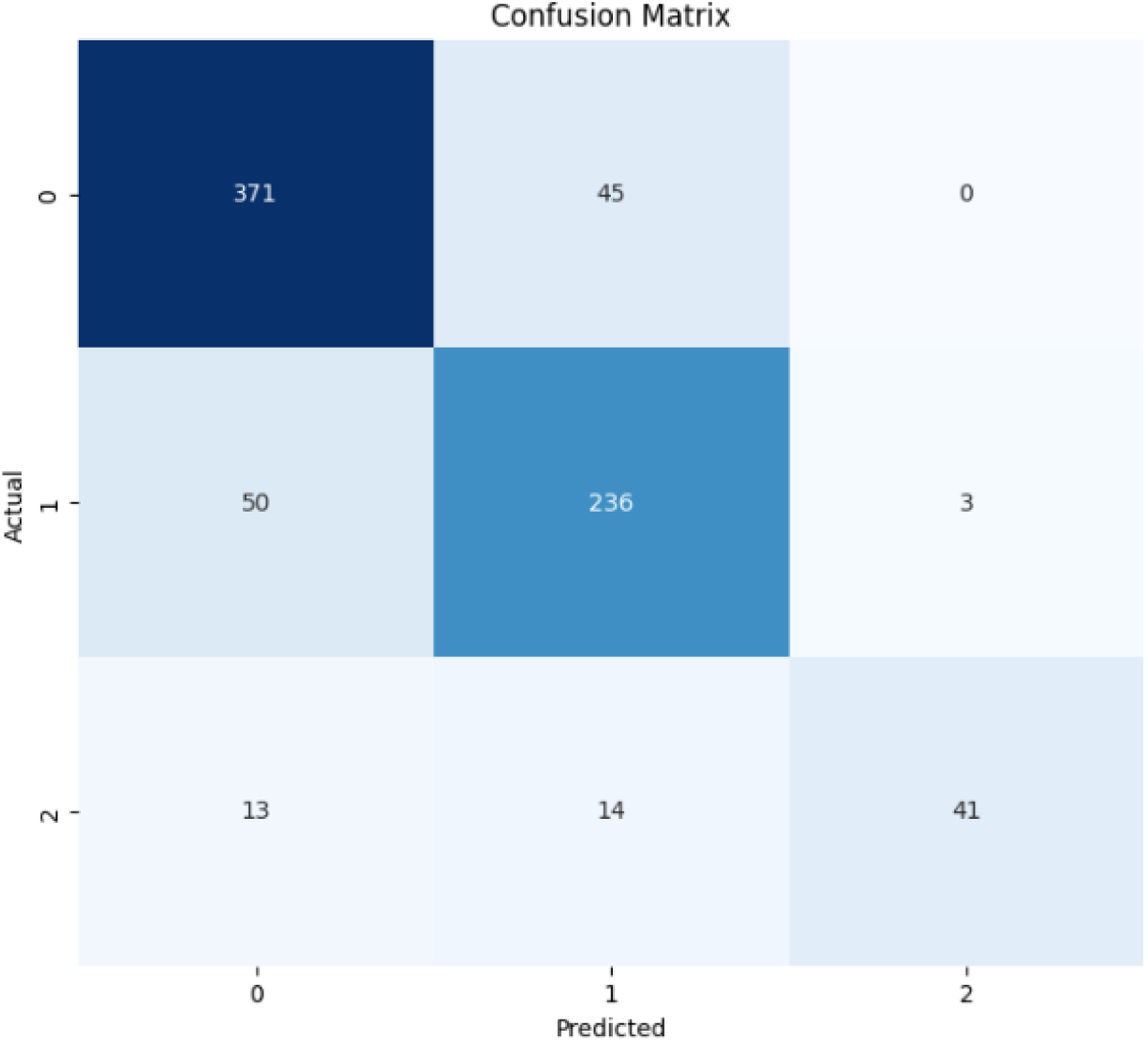
Confusion matrix plot.

The model as shown in figure 14 show good development over epochs with improvements. The training history plots illustrate how the model learns over approximately 120 epochs. In the left plot showing accuracy, both training and validation accuracy rise sharply in the initial epochs, indicating that the model quickly captures key patterns in the data. The training accuracy stabilizes around 0.88–0.90, while the validation accuracy plateaus slightly lower, around 0.83–0.85, suggesting that the model is fitting well but retains a modest gap between training and validation performance. In the right plot, depicting loss curves, both training and validation loss decrease steeply at first and then flatten out, maintaining similar levels from about epoch 10 onward. The close alignment between training and validation loss throughout training suggests that the model generalizes reasonably well and does not suffer from significant overfitting or underfitting. Overall, these plots indicate a stable and effective training process, achieving good performance while maintaining consistency between training and validation results.

**Fig 14:**
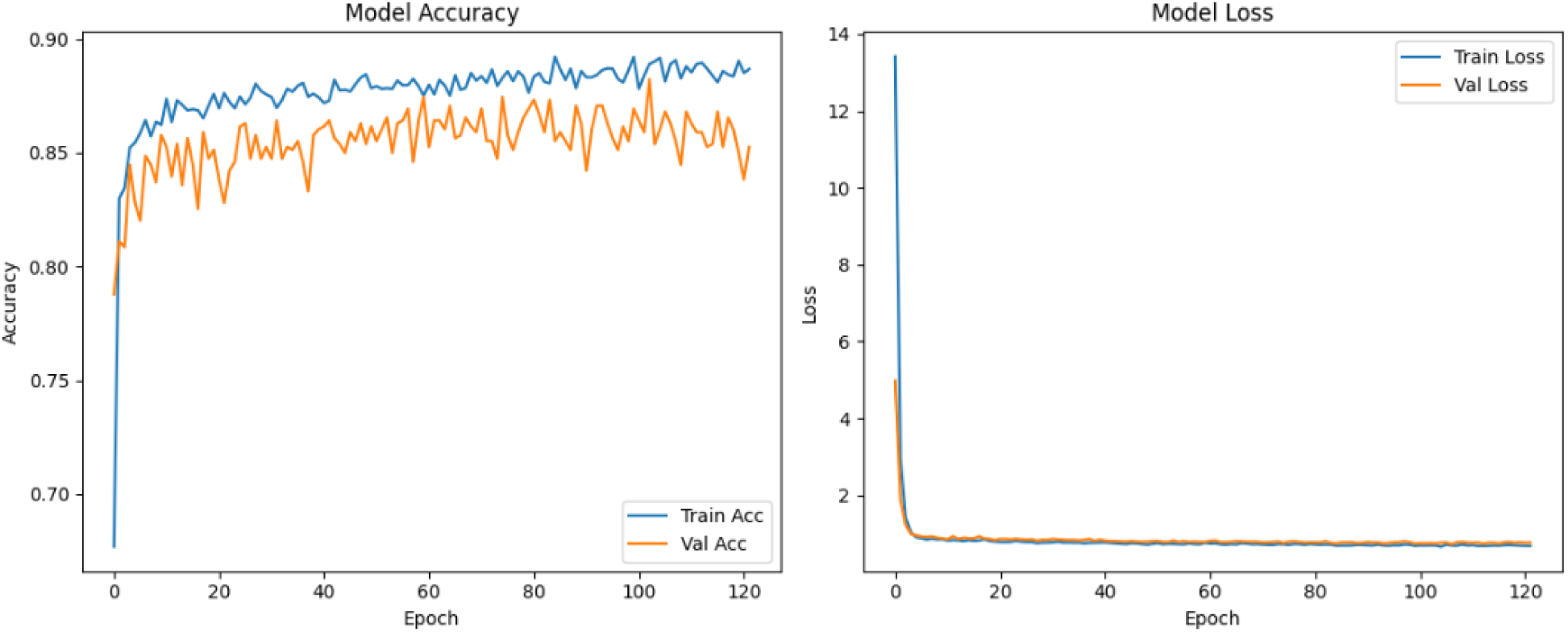
Bi-Directional LSTM GRU training and loss history.

##### f. XGBoost model

The confusion matrix plot in figure 15 is displayed as seen in figure in which for class ‘0’ 403 instances or testing feature set were correctly classified for class ‘0’ similarly, for class ‘1’ 251 instances were correctly classified as class ‘1’ and ‘59’ instances for class ‘2’. The model obtained an accuracy of 100% on training set and Testing Accuracy: 92% accuracy on testing set.

**Fig 15:**
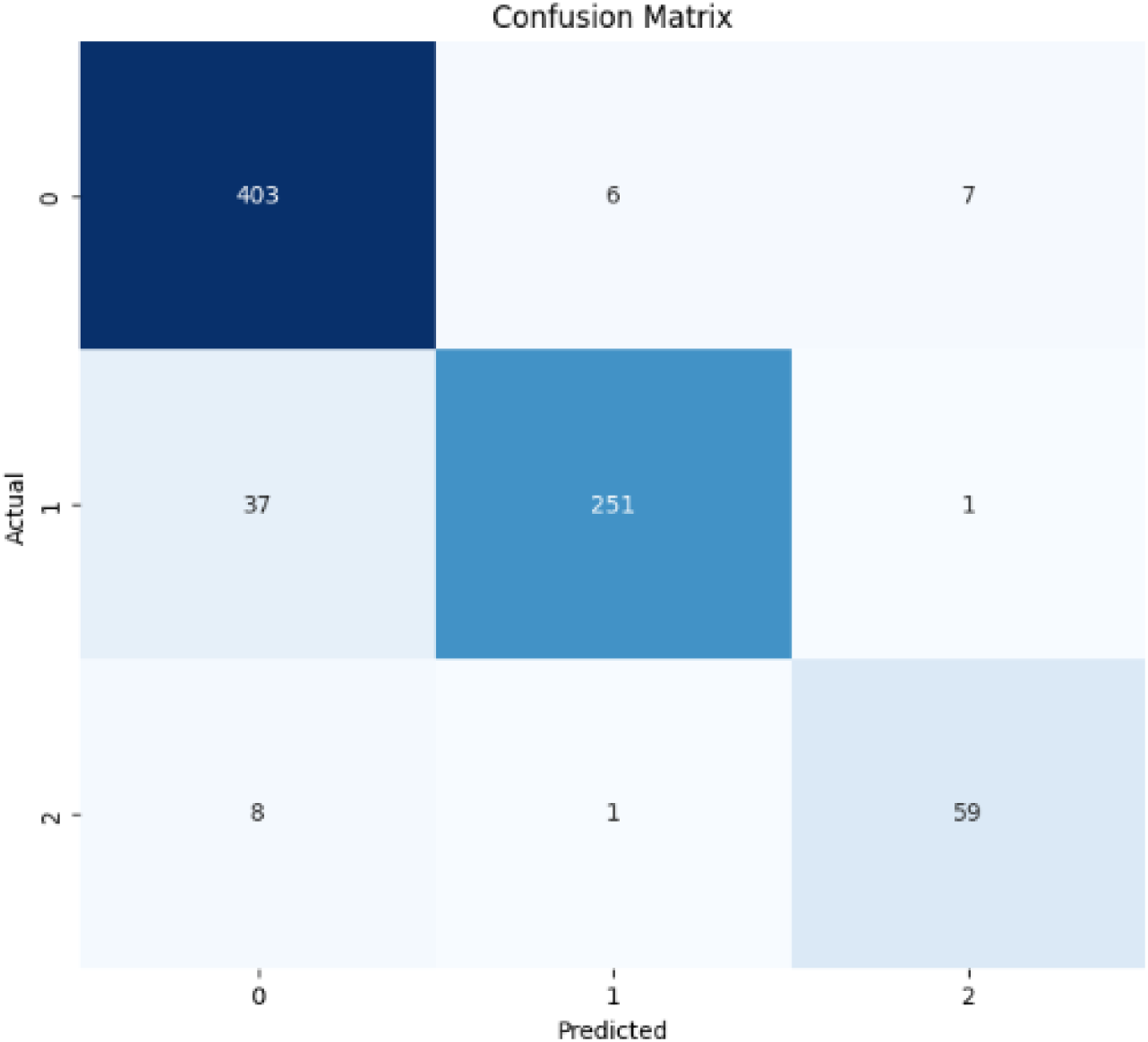
Confusion matrix plot for XGBoost.

Across various models as shown in table 2 and 3, EEGNet achieved the highest testing accuracy of 94%, followed by XGBoost at 92%, Extra Trees Classifier at 90%, and Random Forest at 89%, while Bi-Directional GRU with Attention and Bi-Directional LSTM GRU both attained 84%; further, nested cross-validation confirmed consistent performance, with fold-wise test accuracies ranging from 90.29% to 92.45%, yielding an overall average accuracy of 91.50% and a low standard deviation of 0.0070, indicating the models are stable and generalize well across different data splits. The table 4 shows nested cross validation result for that model.

**Table 2:** Models performances comparison.

| **Model** | **Class** | **Precision** | **Recall** | **F1-Score** | **Support** |
| --- | --- | --- | --- | --- | --- |
| **Bi-Directional GRU with Attention** | 0 (Normal) | 0.84 | 0.91 | 0.87 | 416 |
|  | 1 (CPS) | 0.84 | 0.80 | 0.82 | 289 |
|  | 2 (Electrographic) | 0.88 | 0.63 | 0.74 | 68 |
|  | **Accuracy** |  |  | **0.84** | **773** |
|  | Macro Avg | 0.85 | 0.78 | 0.81 | 773 |
|  | Weighted Avg | 0.84 | 0.84 | 0.84 | 773 |
| **EEGNet stacked model** | Normal | 1.00 | 0.99 | 0.99 | 3479 |
|  | CPS | 0.98 | 0.86 | 0.92 | 2475 |
|  | Electrographic | 1.00 | 0.99 | 0.99 | 682 |
|  | **Accuracy** |  |  | **0.99** | **6906** |
|  | Macro Avg | 1.00 | 1.00 | 1.00 | 6906 |
|  | Weighted Avg | 0.94 | 0.94 | 0.94 | 6906 |
| **Extra Trees Classifier** | Normal | 0.86 | 0.98 | 0.92 | 416 |
|  | CPS | 0.98 | 0.83 | 0.90 | 289 |
|  | Electrographic | 0.92 | 0.68 | 0.78 | 68 |
|  | **Accuracy** |  |  | **0.90** | **773** |
|  | Macro Avg | 0.92 | 0.83 | 0.87 | 773 |
|  | Weighted Avg | 0.91 | 0.90 | 0.90 | 773 |
| **Random Forest** | Normal | 0.85 | 0.97 | 0.91 | 416 |
|  | CPS | 0.96 | 0.85 | 0.90 | 289 |
|  | Electrographic | 0.81 | 0.50 | 0.62 | 68 |
|  | **Accuracy** |  |  | **0.89** | **773** |
|  | Macro Avg | 0.87 | 0.78 | 0.81 | 773 |
|  | Weighted Avg | 0.89 | 0.89 | 0.88 | 773 |
| **Bi-Directional LSTM GRU** | Normal | 0.85 | 0.89 | 0.87 | 416 |
|  | CPS | 0.80 | 0.82 | 0.81 | 289 |
|  | Electrographic | 0.93 | 0.60 | 0.73 | 68 |
|  | **Accuracy** |  |  | **0.84** | **773** |
|  | Macro Avg | 0.86 | 0.77 | 0.80 | 773 |
|  | Weighted Avg | 0.84 | 0.84 | 0.84 | 773 |
| **XGBoost** | Normal | 0.90 | 0.97 | 0.93 | 416 |
|  | CPS | 0.97 | 0.87 | 0.92 | 289 |
|  | Electrographic | 0.88 | 0.87 | 0.87 | 68 |
|  | **Accuracy** |  |  | **0.92** | **773** |
|  | Macro Avg | 0.92 | 0.90 | 0.91 | 773 |
|  | Weighted Avg | 0.93 | 0.92 | 0.92 | 773 |

**Table 3:** Model performances.

| **Model** | **Testing Accuracy** |
| --- | --- |
| Bi-Directional GRU with Attention | 0.84 |
| EEGBoostNet | 0.9588 |
| Extra Trees Classifier | 0.90 |
| Random Forest | 0.89 |
| Bi-Directional LSTM GRU | 0.84 |
| XGBoost | 0.92 |

##### g. SHAP analysis results

The Random Forest feature relevance for each EEG channel, averaged over timepoints, is shown in this bar plot, which shows how much each channel influences the model’s classification choices. EEG signals are high-dimensional and noisy. Using black-box ML models (like XGBoost, Random Forests, or deep networks) can give high accuracy, but we often don’t know which features contribute to seizure detection.

i. SHAP (SHapley Additive exPlanations) assigns each feature a value that represents its contribution to the model output for each prediction.
ii. It’s based on game theory, ensuring fair distribution of “credit” among feature

**Fig 16:**
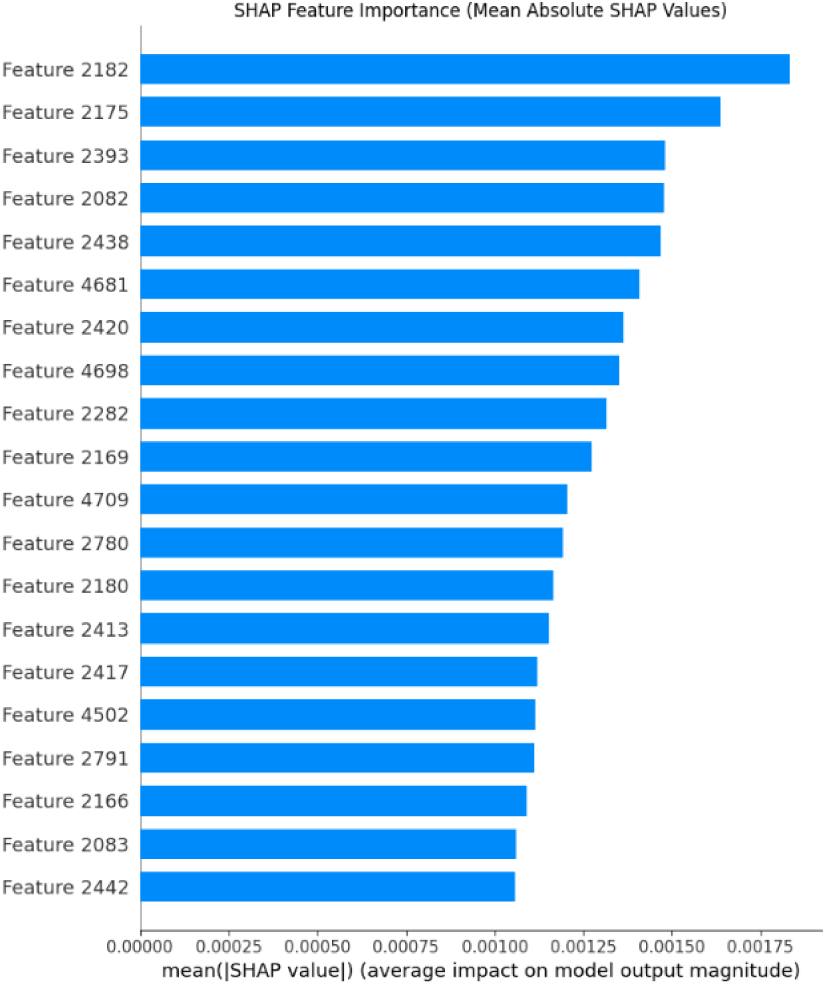
SHAP plot summary.

**Fig 17:**
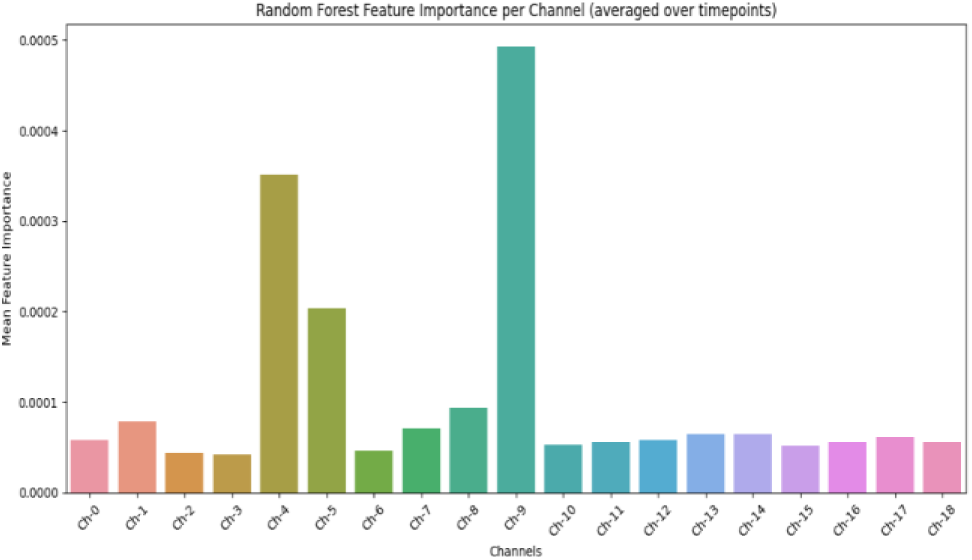
SHAP importance bar plot.

Interestingly, Channel 9 (Ch-9) has the highest mean feature importance, indicating that it contains the most discriminative information for the task. This could indicate that there is a large amount of brain activity associated with the condition under study. While some channels contribute less and cluster closer to zero, others, like Ch-4 and Ch-5, also have very significant relevance. By identifying important brain regions for additional research or focused electrode installation, these insights enhance the interpretability of the model and its potential for clinical use. The figure emphasizes the importance of feature selection in EEG-based machine learning by demonstrating that not all channels are equally useful.

### 3.2 4-classes Predictions

The 4-class predictions are important in dataset as to predict the video-detected seizures too using ML approaches even though samples were very less in comparison to normal requirement to train ML models still this classification is vital to build models that can in future help creating advanced and enhanced algorithm for epileptic seizure classifications of such types.

The traditional ensemble XGBoost model obtained testing accuracy of 91.78 % and training accuracy of 99.82%. Similarly, EEGNet-XGB-RF-ET ensembled, the proposed hybrid network obtained an accuracy of 95.37%. The performance is compared as shown in table 4 and model hyperparameters in table 5.

**Table 4:** Model performance analysis through Nested cross validations.

| **Fold** | **Test Accuracy** |
| --- | --- |
| Fold 1 | 0.9173 |
| Fold 2 | 0.9245 |
| Fold 3 | 0.9167 |
| Fold 4 | 0.9134 |
| Fold 5 | 0.9029 |
| **Average** | **0.9150** |
| **Std Dev** | **0.0070** |

**Table 4:** Classification report for the models.

| Class | Precisi<br>on<br>(XGBo<br>ost) | Recall<br>(XGBo<br>ost) | F1-Score<br>(XGBo<br>ost) | EEGBoostN<br>et | EEGBoostN<br>et | EEGBoostN<br>et |
| --- | --- | --- | --- | --- | --- | --- |
| Normal<br>(0) | 0.89 | 0.96 | 0.93 | 0.95 | 0.97 | 0.96 |
| CPS (1) | 0.96 | 0.86 | 0.91 | 0.96 | 0.93 | 0.94 |
| Electrogr<br>aphic (2) | 0.88 | 0.88 | 0.88 | 0.96 | 0.99 | 0.97 |
| Class 3<br>(3)<br>(Video-detected<br>Seizures) | 1.00 | 0.67 | 0.80 | 1.00 | 1.00 | 1.00 |

**Table 5:** Training Hyperparameters for tree-based models.

| Model | Hyperparameters |
| --- | --- |
| XGBoost | max_depth = 50n_estimators = 1000eval_metric = 'mlogloss'random_state = 42 |
| Random Forest | n_estimators = 100class_weight = 'balanced'random_state = 42 |
| Extra Trees | n_estimators = 300max_depth = 30min_samples_split = 5min_samples_leaf = 2max_features = 'sqrt'bootstrap = Trueclass_weight = 'balanced'random_state = 42n_jobs = -1 |

**Table 6:** Stacked ensembled model hyperparameters.

| Category | EEGNet | Stacked Tree Layer |
| --- | --- | --- |
| Input Shape | (19, 500, 1) | Flattened EEGNet output |
| Feature Scaling | Z-score normalization | StandardScaler |
| CNN Filters | F1 = 8, F2 = 16 | — |
| Kernel Length | 64 | — |
| Depth Multiplier (D) | 2 | — |
| Dropout | EEGNet Blocks = 0.5, FC Layer = 0.25 | — |
| Fully Connected Units | Dense = nb_classes | — |
| Output Activation | Softmax | Softmax via tree-based voting |
| Loss Function | Categorical Crossentropy | mlogloss (XGBoost) |
| Optimizer | Adam | Tree-based |
| Learning Rate | 1e-3 | – |
| Training | Epochs = 200, Batch Size = 32 | – |
| EarlyStopping | Yes | – |
| ReduceLROnPlateau | Yes | – |
| Base Learner | – | Extra Trees |
| Meta Learner | – | XGBoost |

**Table 13:** Hybrid model stratified cross validation performance.

| Fold | Test Accuracy |
| --- | --- |
| 1 | 0.9479 |
| 2 | 0.9276 |
| 3 | 0.9500 |
| 4 | 0.9522 |
| 5 | 0.9428 |
| **Mean** | **0.9441** |
| **Std Dev** | **0.0088** |

Based on the training history plots in figure 18 for the EEGBoostNet model performing 4-class seizure classification, the results demonstrate excellent model performance and training efficiency. The accuracy plot shows that both training and validation accuracy improved steadily throughout the training process, with the training accuracy reaching approximately 98-99% and validation accuracy achieving around 92% by the final epoch. This small gap of about 7% between training and validation accuracy indicates good generalization without significant overfitting.

**Fig 18:**
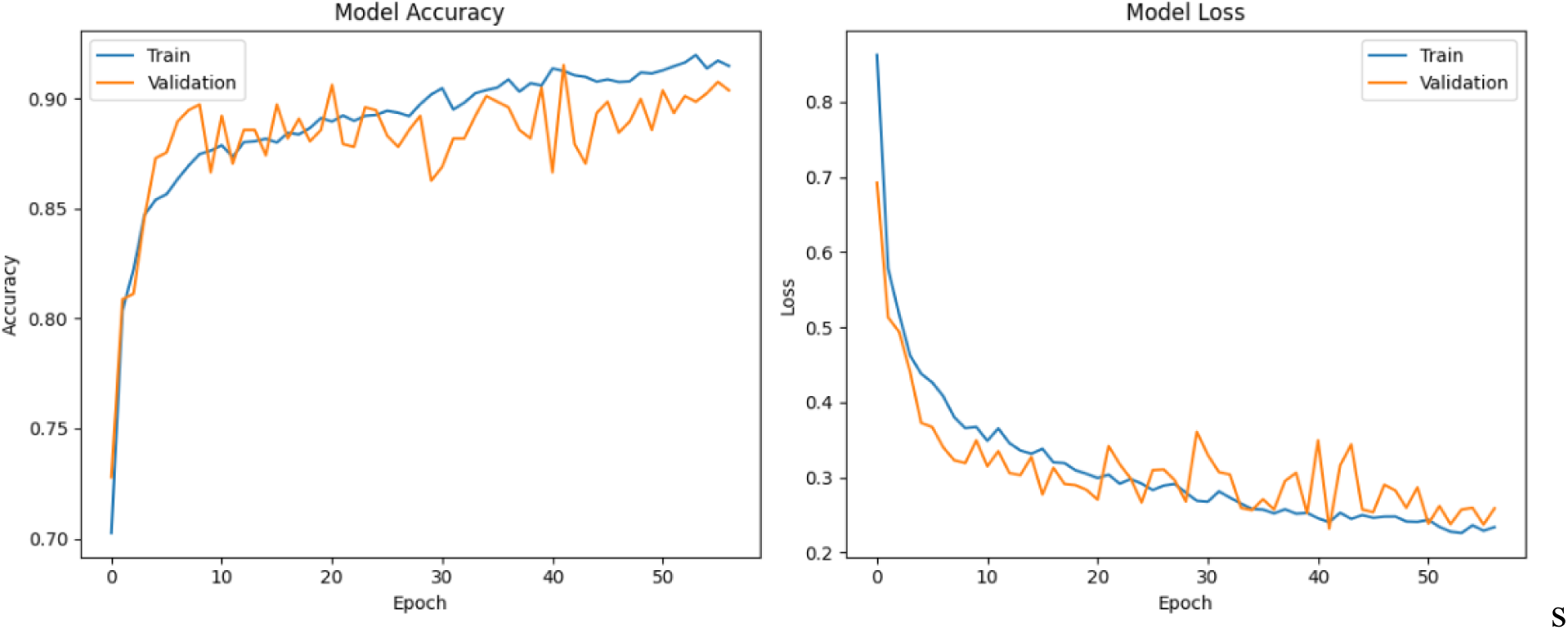
Training and loss history plot for model EEGNet for 4 classes.

The loss plot reveals a similarly positive pattern, with both training and validation losses decreasing rapidly during the initial epochs and stabilizing after approximately 40 epochs. The training loss settled around 0.2 while the validation loss converged to approximately 0.3, maintaining a reasonable margin that further confirms the model’s ability to generalize well to unseen data. The convergence behavior observed in both plots suggests that 80 epochs provided sufficient training time, with most of the learning occurring within the first 40 epochs. Overall, these results indicate that the model architecture successfully learned to distinguish between the three four classes, achieving strong and reliable performance with minimal overfitting concerns. The confusion matrix plot for model is depicted in figure 19.

**Fig 19:**
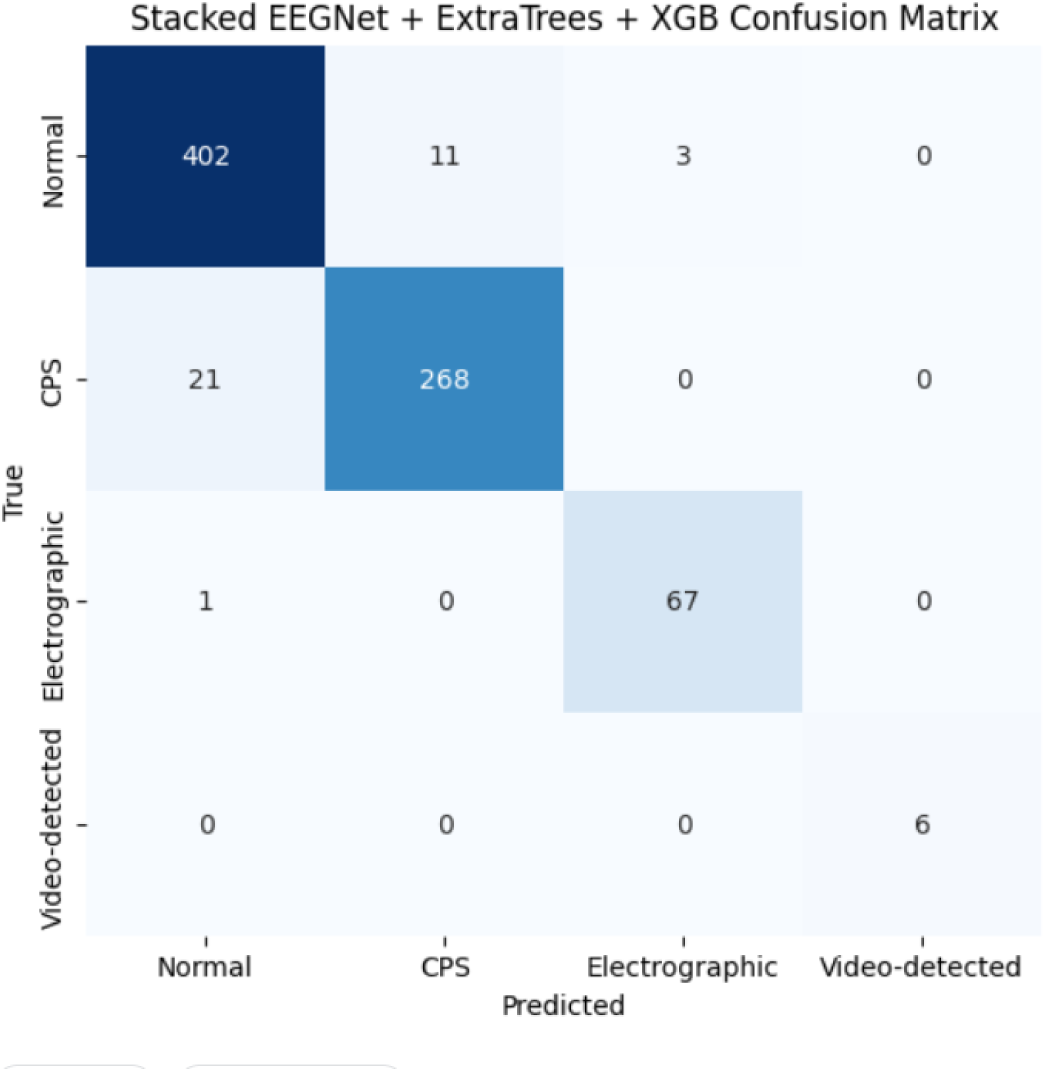
Stacked EEGBoostNet confusion matrix plot.

The model performance history plot is shown in figure 18. The bar chart displays the average feature importance of different EEG channels (Ch-0 to Ch-18) as computed by the Random Forest model. Each bar represents how much a particular channel contributes to the model’s predictions on average across timepoints. Among all channels, Ch-9 stands out significantly, showing the highest mean feature importance by a large margin. This indicates that Ch-9 carries the most critical information for distinguishing between classes in the seizure detection task and SHAP values also quantify how strongly each feature influences model predictions. Therefore, this confirm that Ch-9 has the greatest impact on model performance, suggesting it may capture key patterns or signals relevant to epileptic activity in the EEG data.

With this system early warning can be triggered, and early alert notification can be sent via IoT on email. This system aids in automating appliances using microcomputer-based processing of the EEG signals from the victim so that when the affecting onset is predicted by the model, the patient room conditions can be automated to open a bulb, fan, or similar device using relay-based triggering. In summary, this system acts as feedback for making an IoMT (Internet of Medical Things) home automation for seizure victim aid and managing favorable conditions to provide a relief aspect to the victim. Thus, patients can be aided easily with the help of the early warning and notification system along with the control mechanism for various devices in the home environment.

### 3.3 Limitations and Future works

Thus, the proposed approach shows a significant concept in EEG signal classification for detecting various kinds of seizures and is vital to identify their impacting features, the channels that much affect the prediction, as well as choosing the best model for performance enhancement. With current work, the main drawback is the lack of enough training data for class 3 (videographic seizures), and for that purpose, in the future, such data need to be included with higher volume. The models developed shall be enhanced, as still 95-96% accuracy isn’t best suited for real-time deployment for purposes of IoMT ecosystem development.

As proposed work has depicted an IoMT concept for aiding patients, a real-time evaluation couldn’t be made due to a lack of EEG headsets and more advanced devices. In the future, rigorous testing for such prototyping can be carried out.

## 4. Conclusion

To conclude, an ML model like EEGBoostNet ensembled with tree models with newer architecture has a higher potential to gather greater performance for EEG results with approximately 96% accuracy for seizure detection. The models trained on the dataset show that prior to various epileptic conditions, early warning can be triggered using ML-based detection via victim EEG data. The EEG preprocessing and normalization applied allowed the hybrid-deep learning models to perform and learn patterns very well. This approach can be useful for deployment in real-time prototyping for IoMT edge and medical device development. With this approach, future intelligent biomedical systems based on EEG can be enhanced.

## Data Availability

data is publicly available

https://data.mendeley.com/datasets/5pc2j46cbc/1

